# Hospital based autonomous pre-clinical screening of COVID-19: An emergency triage using a vital signs recording system, Paris-Ile de France region

**DOI:** 10.1101/2020.06.17.20133934

**Authors:** Albert Brizio, Valérie Faure, Franck Baudino, Arnaud Wilmet, Jean-Paul Gonzalez

## Abstract

The COVID-19 pandemic, has had a dramatic toll worldwide on the populations but also has been essentially supported by the existing public health system, particularly hospital-based emergency wards and intensive care units.

In France, the first cases were identified on the 24^th^ of January 2020. The first epidemic sprout emerged in the Eastern part of the country and spread in two weeks towards the center to the Paris-region where it peaked on the 14^th^ of April 2020. In Paris and the region around it, the intensity of the epidemic has increased significantly to have a strong impact on all public and private hospital systems in a few weeks.

During France’s 2020 COVID-19 epidemic, a private hospital went into a major organizational change of its Emergency Department which mainly included the use of a Telemedicine Booth for vitals automatic recording during triage procedures.

The present study describes the organizational scheme adopted by the hospital and discusses the data of 1,844 patients that attended the facility over a month. While among them, 766 patients were engaged in a automated triage process supported by a Telemedicine Booth. Patients’ clinical characteristics are comparable to those found in international literature during the COVID-19 pandemic.

The use of the Telemedicine Booth as a screening process facilitated patients’ flow. It usefully participated in the patient rapid orientation, relieving the hospital emergency department, actively contributes in a safe and secure environment highly trusted by the hospital staff and health workers. To our nowledge, the Telemedicine Booth use as a screening process during an epidemic constitutes the first contribution to such an innovative approach.

## INTRODUCTION

The World Health Organization (WHO) declared, on March 11^th^ 2020, the Coronavirus disease 2019 (COVID-19) as a pandemic, pointing to the increasing number of infections among more than 110 countries and territories worldwide, and the risk of further global spread. Considered by WHO, Europe had become by that time the epicenter of the pandemic, the level three of the COVID epidemic emergency was officially declared by the French authorities on March 14^th^ 2020^1^.

According to the local governmental instructions, the Vert Galant Hospital (Fig. 1), as a private health facility within Paris metropolitan area, shifted from normal activity to a crisis management on March 19^th^. The newly developed engagement involved the whole hospital aiming principally to facilitate the influx of new patients and avoid overwhelming the potential of the Emergency Department (ED) (i.e. inclusive of the Intensive Care Unit). This article describes, analyzes this innovative organizational scheme and discusses the outcomes of its development and the impact over a month’s period of time (March 19^th^ to April 19^th^, 2020) during the intense inception of COVID-19 epidemic within Paris metropolitan area.

**Figure 1:**
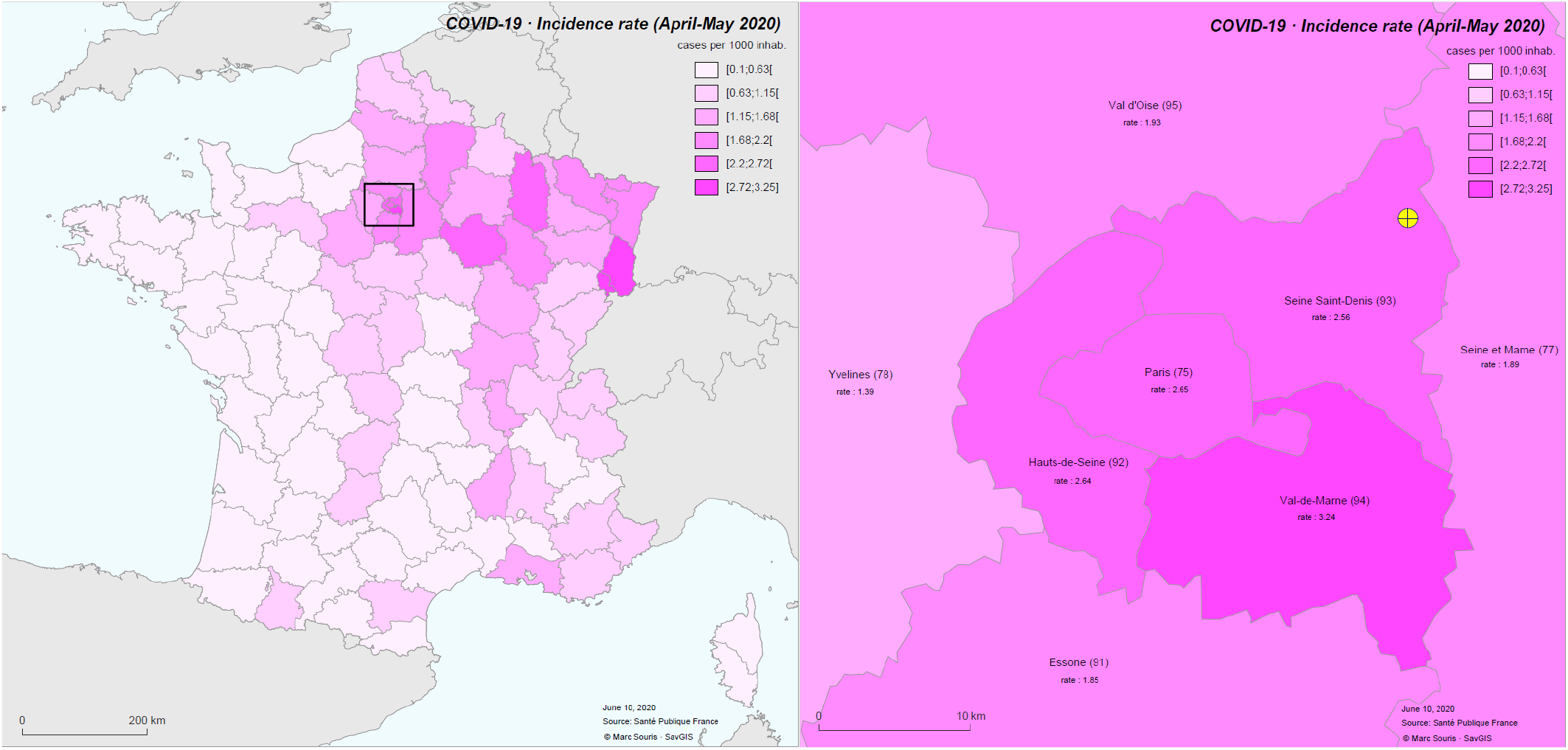
Incidence of COVID-19 cases and localization of Paris and the Vert Galant Hospital in the Seine Saint Denis Departement (modified from Santé publique France)

## PERSONNEL, METHOD, AND MATERIAL

### Study site

The Vert Galant Hospital is a 100 bed private hospital with an intensive care unit (ICU) of 14 beds of an emergency department (ED) with a potential to manage a mean flux of hundred patients a day, as well as other medical wards including: gynecology and obstetrics; orthopedics; oncology; cardiology; gastroenterology; pulmonology; urology; and endocrinology. It essentially covers a low-medium income population of the Seine Saint-Denis, 93 Department (French District Unit), wich is one of lowest income areas in France with a poverty rate of 26,9% as compared to the 14,3% of continental France^2^. The hospital accepts all patient with national health insurance as well as irregular migrants without insurance.

The Vert Galant emergency department was designated by its hospital board to manage the increasing flow of patients due to the intensive rise of the COVID-19 epidemic. Therefore, by setting up a new organizational scheme, measures were taken to lessen risks for the health workers without reducing the efficiency of patient care.

### Telemedicine Booth

One of the main measures of the new hospital strategy during the epidemic crisis was to implement the use of a Telemedicine Booth (H4D ConsultStation®) which allow, among other health data collection, the measurement and the automatic recording of the patients’ essential vital signs. The Telemedicine Booth is also a teleconsultation facility allowing, besides vital sign collection, an interactive video conferencing with a medical practitionner when needed, the latter was not used in the present context of the management. The Telemedicine Booth is a Class 2 certified medical device and can be described as a fiberglass cylinder with a seat, a video and audio system plus a touch video screen for patient-machine interaction, printer, and a series vital signs’ measurement devices (Table 1).

**Tab 1:**
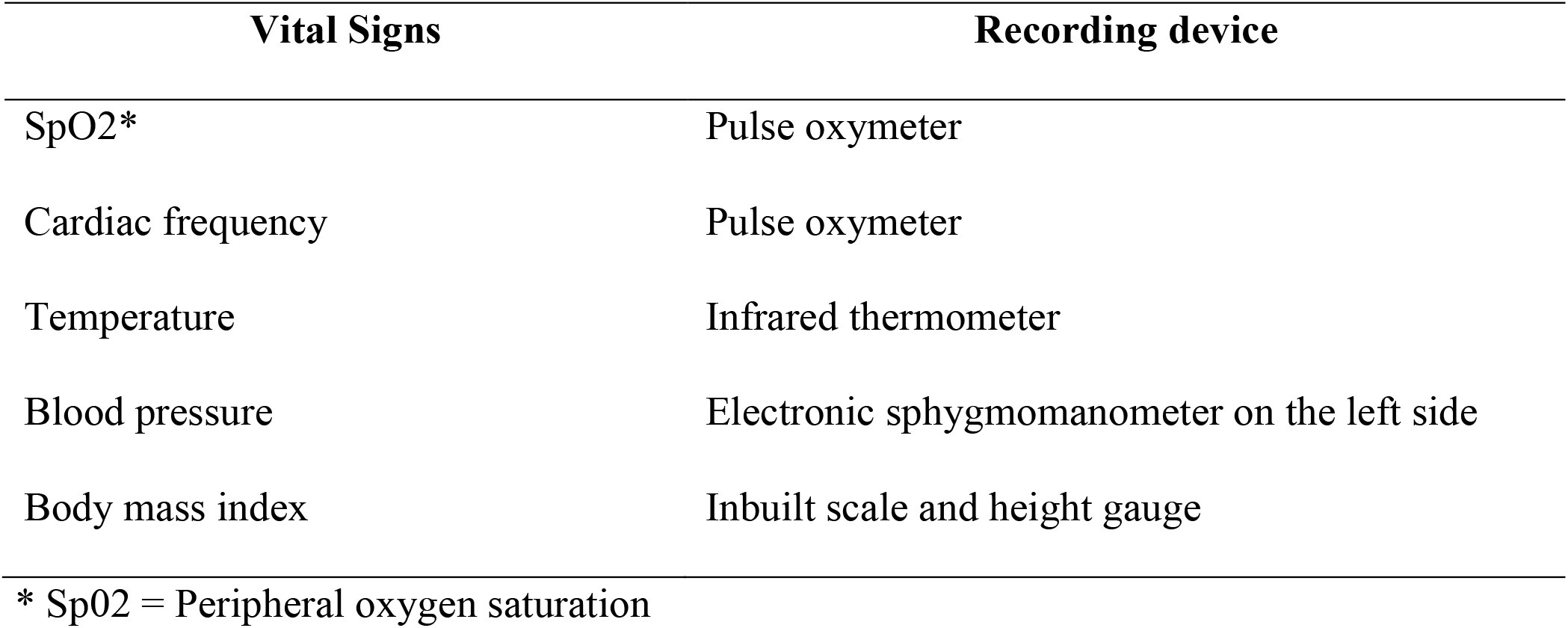
The vitals recorded by the hospital based Telemedicine Booth during the COVID-19 epidemic in the Paris metropolitan area (may-April 2020)

### Health workers and administrative staff

The ED, on a daily basis, is staffed by two (2) senior medical practitioners (MP), three (3) nurses, three (3) aids/stretcher-bearers, two (2) secretaries, and during the night shift by one MP (1), two (2) nurses, one aid/stretcher-bearers, and one (1) secretary.

### Management and organizational scheme

The ED management was and is under the responsibility of the MP and the director of the hospital. According to emergency plan, a COVID-19 crisis unit was set up while routine hospital activity was reduced or suspended for some non-essental sectors. Two-thirds of the surgery wards were transformed in COVID-19 wards and the number of ICU beds was increased by two fold (i.e. 28 beds). One of the ED MP was mandated as coordinator in charge of the organizational change and supported by the chief nurse. Technical choices were team verdicts involving the ED MP, the Chief Nurse, and a crisis unit representative.

The COVID-19 ED system was set up for the time being, it was designed to focus on the implementation, to follow existing recommendations as far as possible as well as previous experiences. Records were kept of all activities mainly using existing procedures. Ultimately, the principal objectives where: 1/ To create a specific Covid-19 patient flow in the existing facilities; 2/ To maintain a separate non-COVID-19 flow; 3/ To manage the patient and record their vital signs in view of reducing infectious risk (i.e. infection control) and protecting the health workers and the patients. In order to achieve these objectives, the capacity of the ICU increased by two-fold, and patients’ flow was redesigned. The outpatient activity and facilities were canceled, such facilites, situated in the immediate proximity of the ED, were designated for non-COVID-19 emergency space. Also, it was decided to keep as the single public entrance to the hospital, the ED entrance, and to create an external temporary initial triage station in order to separate potential COVID-19 from non-COVID-19 patients before entering the facility. The parking area, in front of the ED entrance, was utilized to set up a tent (fig.1).

### Strategic engagement

Triage the act of prioritizing patients’ access and determining severity, usually performed by a trained nurse, was considered strategical. The pre-existing ED triage room was small, with open entry with the secretaries’ room. To avoid any risk of transmission for nurses during the triage, it was decided to use a Telemedicine Booth allowing an autonomous recording of the vital signs in association with a questionnaire. Human supervision was considered essential to speed up the triage process and to provide the necessary help if needed. It was decided to install the Telemedicine Booth in the ED waiting room and train a group of stretcher-bearers for this novel role. Health workers received two hour training focusing on the COVID-19 infection risk including: The use of personal protective equipment; the technical management of the Telemedicine Booth; infection control (hands cleaning, Telemedicine Booth disinfection and safe disposal); the recognition of respiratory distress as well as other potentially severe conditions.

The triage itself was modified during daily activity by replacing the FRENCH 2018 triage grid^3^ for non-COVID-19 patients maintained in the blue sector, by a three steps procedure for COVID-19 patients inclduing: A first step at the tent, a questionnaire was intended to identify COVID-19 patients; a second step, in the red sector, the vital signs taken at the Telemedicine Booth intended to first evaluate the severity of clinical presentation; and a third step for the nurse to re-evaluate the syndrome severity while contextualizing it with the help of medical history before the medical examination by the MP.

During night hours the organization shifted while the night attendance rate did not justify the Telemedicine Booth facilitator’s use and did not include it for screening.

### Patient pathway

Outside of the hospital premises, the parking access was controlled, while immediately front of the entrance the “first triage” tent was set up. A medical student or a nurse fully dressed with personal protective equipment (PPE) sat at the entrance for: Guiding the patient to disinfect his hands with hydroalcoholic gel and to accurately fit a provided surgical mask; applying a questionary on symptoms and reasons to attend. The questionary was based on available clinical guidelines and a formal list of symptoms to identify suspected COVID -19 cases^4^.

Patients waited in a single row at 1,5m distance one to each other following marks on the ground. Once oriented to his destination, the patient was invited to cross the tent and enter the building through a glass sliding door. The pathway used by each patient was directed through a color-coded system (i.e. red Covid-19 suspected; blue non-Covid-19) from the time he entered the hospital to the time he left the triage and was redirected based on the data acquired (Fig. 2). Non-COVID-19 patients followed blue signs leading to blue doors giving access to the “blue” sector. The “blue” patients would simply proceed straightforward, enter the blue zone, and be attended. When needed, they would attend the blue sector of radiology, if dismissed, would exit through a dedicated door. COVID-19 suspected patients would follow the red signs leading to the “red” ED sector. The “red” patients, had to turn left, go through a transparent curtain, then an automatic sliding door leading to the waiting room, before being welcomed by the stretcher-bearer fully dressed in the appropriate PPE and in charge of the Telemedicine Booth. In the autonomous “check-up” modality, the patient was guided through the Telemedicine Booth by a video and audio tutorial. At the end of the process, the Telemedicine Booth print out a report. This process took usualy seven (7) minutes to the patient with no external help.

**Fig 2:**
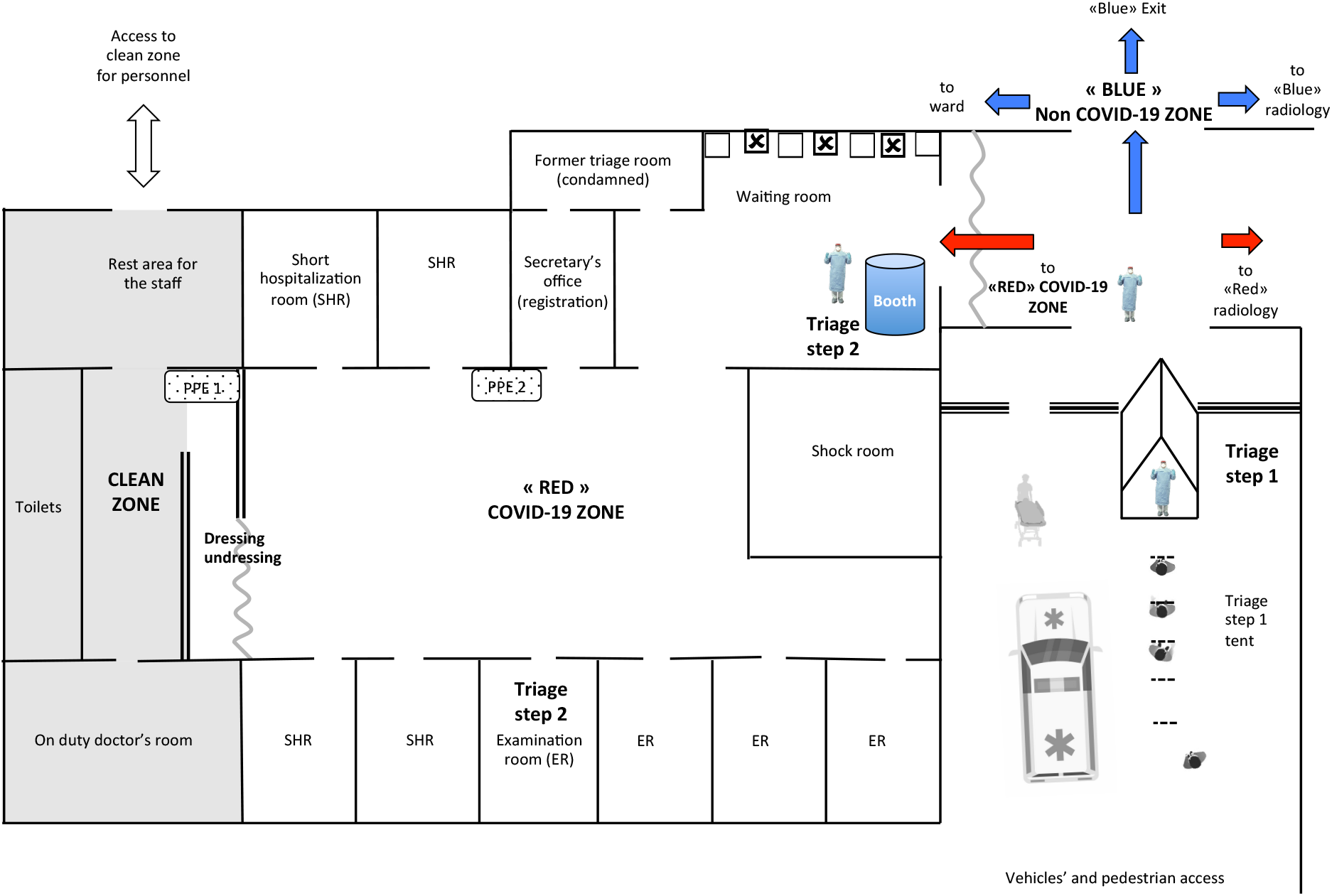
COVID-19 patient pathway upon admission to the Emergency Department of the hospital during the epidemic of Paris Metropolitan area (March-May 2020).

Patients transported by ambulance were bypassing the tent and entering directly to the dedicated wards. During the night, the first triage was maintained in the tent although in the red sector, the Telemedicine Booth was bypassed, and, once registered the patient was directly admitted to the examination room where the vital signs were measured by the nurse. In order to maintain security distances one out of two seats of the waiting room was marked with tape crosses for not to be used. Once the administrative formalities finished at the registration desk, the patient was invited to enter the Telemedicine Booth, having first disinfected his hands. After the vital signs recording, the patient stayed in the waiting room under the supervision of the specially trained stretcher-bearer unless the saturation level was below 95%. This would lead to immediate admittance of the medical examination. Once admitted to the examination room, a nurse took the patient’s personal history, measured oxygen saturation and respiratory rate. Followed the medical examination. If required, the patient was accompanied or redirected to the COVID-19 dedicated CT-SCAN (Computed tomography Scanner) for lung examination following a specific “red” path.

The health worker entered from a separate entrance in the “clean” part of the ward. A buffer zone, called dressing/undressing zone, separates it from the red zone. In the clean zone, the health worker dressed with PPE before entering the red one. During their activity in the red zone, overcoats, aprons, gloves, and masks, replacements were available. While returning in the clean zone, undressing was mandatory as was the disinfection of the hands following infection control guidelines. The red and blue sectors initially had two seperate teams, but organizational needs led to frequent shifts of health workers from one team to the other. Except for MP, two separate health worker groups covered the 12 hours day and night shifts.

### Activities evaluation

Evaluation procedures were not specifically included in the project design. The register at the tent, the patient management software, and the Telemedicine Booth activity software ensured data collection with accuracy. All software complied with existing French regulations and laws. An *a posteriori* survey was conducted among the personnel of the ED for qualitative feedback including a questionary concerning their perception of workload, contagious risk, and difficulties encountered with the Telemedicine Booth were submitted.

## RESULTS

### Quantitative indicators of activity

Over the study period, which corresponds to the epidemiological peak of COVID-19 in France^5^, 1,844 patients attended the Hopital du Vert Galant ED. 766 of them (41.5%) were triaged at the tent as suspected COVID-19 cases and were directed to the “red” sector (RS). The total number of admissons in the RS is higher (838) due to a 9.3 % readmissions. The population’s mean age was 47.6 years (46.0 yo female, 49.0 yo male). 61% (51.9 % males) of the patients were diagnosed as COVID-19 positive, either clinically or with the support of RT-CPR and Chest CT-SCAN (Computed Tomography Scanner). Out of the 469 diagnosed COVID-19 positive, 50.9% had a diagnostic confirmation by RT-CPR and/or Chest CT-SCAN. 13.4% of the total screened patients were hospitalized (among them 67% males) including 2.5% in ICU (70% males).

88% attended the ED between 8 AM and 8 PM and, among them, 68% passed through the Telemedicine Booth for autonomous vital signs recording. The data collected from 47 patients allowed to calculate the mean time between the first triage at the tent and the registration by the secretary (15,6 minutes). The registration to Telemedicine Booth time was 12,1 minutes. The mean time spent by the patients in the Telemedicine Booth, calculated for all patients, lasted for only 4,2 minutes (Table 2).

**Table 2:**
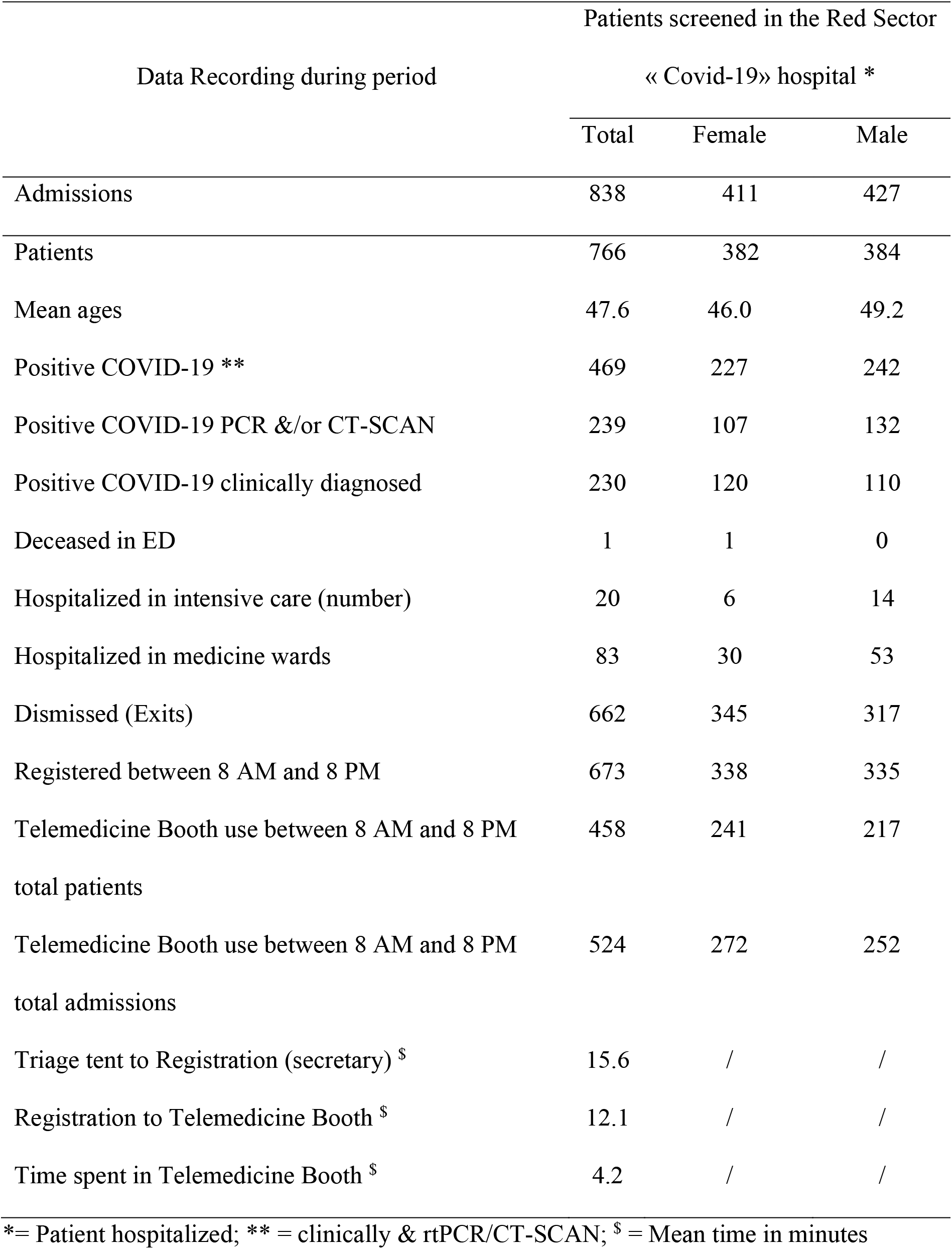
Main quantitative data acquired during the during the COVID-19 epidemic in the Paris metropolitan area (may-April 2020)

The new organization set up took three days and needed the workload of ten (10) health workers. 21 full-time nurses and nurse-assistants and, 31 individual contractors working for the ED during this period.

Out of the 21 full-time health workers, 15 answered to the survey evaluating their subjective perception of the new organizational scheme: 7 found their workload increased, 4 unchanged, and 4 reduced, while 12 did not find difficulties related to the use of the Telemedicine Booth and 9 felt that the use of the booth increased their safety. Among the full-time health workers, 2 only tested positive by RT-CPR without proven origin (domestic or profesional) of the infection.

## DISCUSSION

In comparison to the same time period in 2019, at the Vert Galant Hopital, there has been a 43.0% reduction of the ED’s attendance. This reflects a global phenomenon during the COVID-19 crises^6^ while unexpectedly, the COVID-19 suspected patients were less than the half of the usual frequentation, nevertheless, it was about 6 times higher than the COVID-19 attendance rate of the 93 Department’s public hospitals (41,5% % versus 6,8%)^7^. This could be explained due to close-by population considering the hospital as a “COVID hospital” refraining them to attend the hospital for other medical reasons. Altogether, these figures show the importance of maintaining adequate services for non-COVID-19 cases or any concurent critical epidemic.

The risk of focusing clinical inquiry on COVID-19 symptoms concerned mainly the patients presenting atypical symptoms of the disease and false positives. Therefore, the initial triage not only has to avoid under-triage for the contagious disease but also under-triage of the other concurent conditions not related to it. Possibly, the multi step triage that has been used during the study, appears to be the most appropriate approach to avoid such risk. By assuming the accuracy of clinical diagnosis, an over-triage of 39% was estimated. Also, an over-triage of 25 to 50%^8^ is accepted for traumas, the present over-triage may be acceptable for effectively reducing the potential risk of COVID-19 positive patients admittance to non-COVID-19 areas. However, over triage in epidemic conditions exposes false positive patients to infectious risk. With this perspective, the 39% figure may be considered excessive but limited data are available on this matter from the medical litterature.

The 13.4% COVID-19 hospitalization rate is half of the one registered by the department’s public hospitals (27,2%)^9^ and this could be partially explained by the relatively young age of the population who have attended the Vert Galant hospital facilities.

Although males represented half of the COVID-19 positive patients, they denote 67% of the one hospitalized and 70 % of the one ICU admitted. This is in agreeemnt with other produced elsewere of an increased occurrence of severe COVID-19 cases incidence in males^10^.

Moderate underuse of the Telemedicine Booth is probably due to the speeding-up of the flow. However, to avoid any delay, when the Telemedicine Booth was in use and an examination room was available, the patient was directly admitted to the room thus bypassing the Telemedicine Booth. Nonetheless, about 35 nurse-time hours were spared over a one month period.

The time between registration and the vital signs taking by the Telemedicine Booth is about nine (9) minutes less than the time between registration and vitals measurement by the nurse in normal times (22 minutes). The average total time from the gate triage to the Telemedicine Booth was 27.7 minutes. The mean vital signs’ recording time at the Telemedicine Booth was 61%, less than the 7 minutes needed for a complete autonomous video-guided vital signs recording (used for check-up in a non-epidemic context). This difference is due to the dedicated personnel that actively assisted patients and often skipped the video tutorial used for patient self-guidance. Generally speaking, patient’s security was enhanced by the new system by reducing the vital signs recording time and ultimately improved patient’s supervision.

Despite the consistent decrease of patients’ attendance, the majority of the personnel did not perceive their workload as reduced because the organizational constraints linked to the new triage, dressing, disinfection procedures, and patients’ anxiety. The majority did not consider the Telemedicine Booth as a burden when more than half of them considered that its use increased their own safety. If a few expressed some concern, it was always associated with human error.

The incidence of COVID-19 among non-medical health workers can be evaluated with difficulty because of the small sample size and the numerous confounding factors: shifts between red and blue zones, the absence of routine testing, and the absence of a baseline. To our knowledge, statistical data concerning health workers being not yet available for France, the 14% of symptomatic COVID 19 health workers found in this study (beside a non-proven infection origin) seems in agreement with the 10% infection rate among nurses found in Lombardy, Italy^11^.

The importance to adjust the facility to the epidemic context and to protect the health workers from infection was used as an opportunity for the patient’s health education. Therefore, necessary time was taken for patients’ guidance, in particular to accurately practice hands disinfection and masks fitting upon admission for triage at the tent and Telemedicine Booth use.

## CONCLUSION

This case study illustrates how a major change in organizational habits centering on the introduction of an automatic vital signs recorder in an emergency department can very well possible with a limited disadvantage. The presence of the Telemedicine Booth did not slow down the patients’ triage process, and potentially improved it by an efficient triage and a better trust of the health worker adherence to protective measures.

Although, more research would be needed to fully identify the pros and cons induced by the use of automated vital signs recording Telemedicine Booth during an epidemic in hospital settings. This experience constitutes a first contribution to the exploration of this innovative approach.

## Data Availability

All data are available at the Hopital Prive du Vert Galant, Tremblay en France, France.

## Acknowledgments

This work and the study behind has been possible with the exceptional support of RAMSAY SANTE, Health for Development and the participation of the staff of the Vert Galant Hospital, Madame Sandrine Legras, Dr. François-Xavier Peccaud, and Dr. Florian Gontier. We want to thank Ms. Soukaina Gonzalez for writing assistance, technical editing and language editing.

## Authors Declarations

The authors confirm that all relevant ethical guidelines have been followed, and any necessary IRB and/or ethics committee approvals have been obtained.

The details of the IRB/oversight body that provided approval or exemption for the research described are given below: NA

All necessary patient/participant consent has been obtained and the appropriate institutional forms have been archived.

The authors understand that all clinical trials and any other prospective interventional studies must be registered with an ICMJE-approved registry, such as ClinicalTrials.gov. I confirm that any such study reported in the manuscript has been registered and the trial registration ID is provided (note: if posting a prospective study registered retrospectively, please provide a statement in the trial ID field explaining why the study was not registered in advance). The authors have followed all appropriate research reporting guidelines and uploaded the relevant EQUATOR Network research reporting checklist(s) and other pertinent material as supplementary files, if applicable.

## Legend

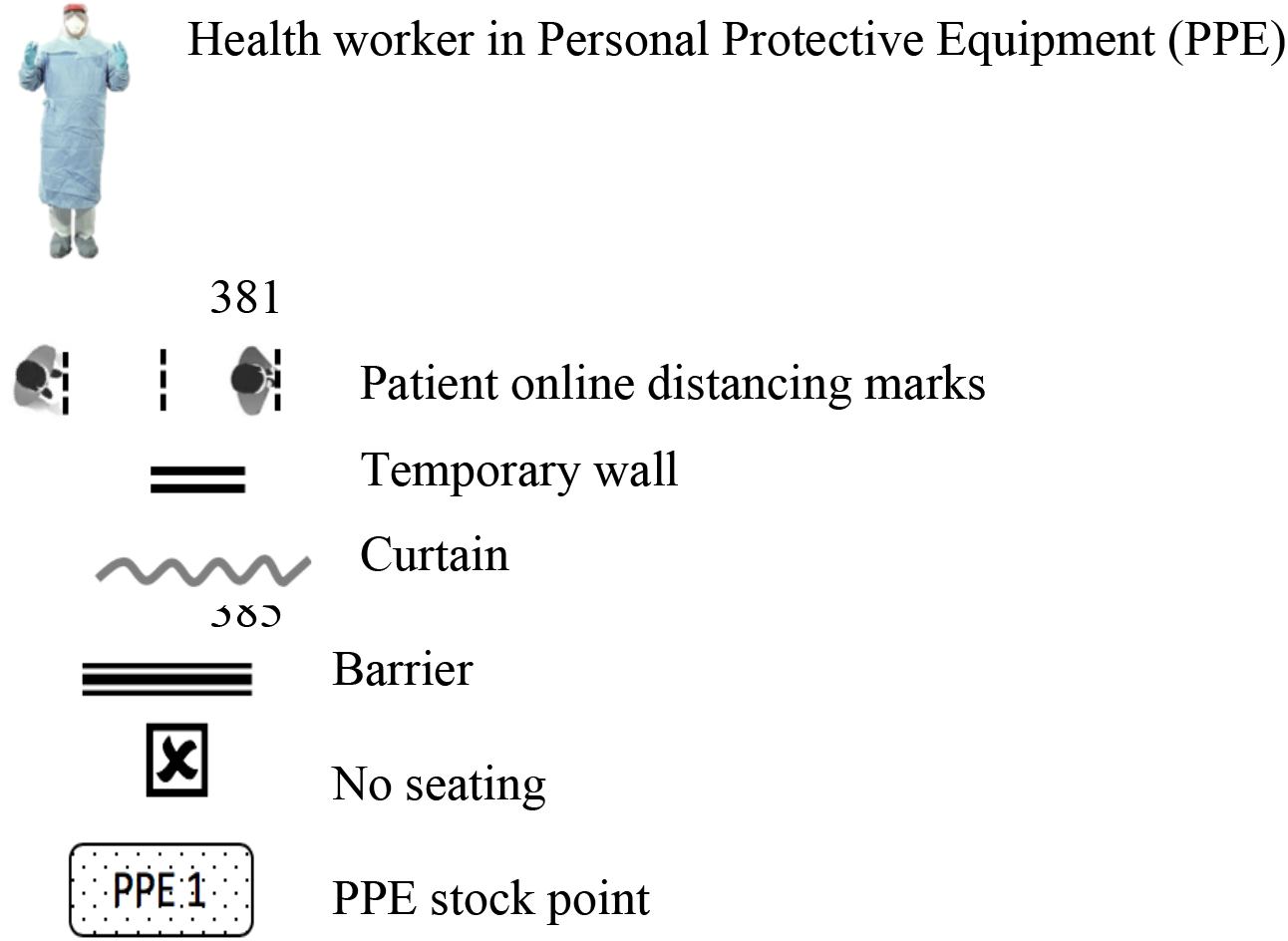

